# Is there a resistance-threshold for macrolide consumption? Positive evidence from an ecological analysis of resistance data from *Streptococcus pneumoniae, Treponema pallidum* and *Mycoplasma genitalium*

**DOI:** 10.1101/2020.10.13.20212043

**Authors:** Chris Kenyon, Sheeba S. Manoharan-Basil, Christophe Van Dijck

## Abstract

**Background:** If we were to keep macrolide consumption below a certain threshold, would this reduce the probability of macrolide resistance emerging? No study that we are aware of has addressed this question.

**Methods:** We assessed at a country level if there was a macrolide consumption threshold for the selection of a prevalence of macrolide resistance of over 5% in *Streptococcus pneumoniae, Treponema pallidum* and *Mycoplasma genitalium*.

**Results:** We found evidence for a macrolide consumption threshold of 1.3 defined daily doses per 1000 inhabitants per day (DID) for *M. genitalium*, 1.8 DID for *T. pallidum* and 2.3 DID for *S. pneumoniae*.

**Conclusions:** Our results provide further motivation for macrolide stewardship campaigns that strive to reduce macrolide consumption to levels below at least 2 DID.

## Background

Whilst it is widely accepted that antimicrobial consumption plays a crucial role in the emergence and spread of antimicrobial resistance (AMR), there is little consensus as to whether or not this association is linear (1). Certain authors have claimed that there is a ‘use it and loose it’ relationship whereby AMR is viewed as the inevitable outcome of using antimicrobials (1, 2). In contrast, Levy hypothesized that there were thresholds of antimicrobial consumption, below which AMR was unlikely to emerge and spread (3). A recent longitudinal study has provided epidemiological evidence to back up the existence of such thresholds for a range of bug-drug combinations. In this study, minimum antimicrobial consumption thresholds were identified for carbapenem resistance in *Acinetobacter baumannii*, cephalosporin resistance in *Escherichia coli*, gentamicin resistance in *Pseudomonas aeruginosa* and methicillin resistance in *Staphylococcus aureus* (1). This study was, however, limited to isolates from inpatients in 5 European countries. It would be useful to know if there were also thresholds of antimicrobial consumption in the general population. If so, these could be used as targets in antimicrobial stewardship campaigns (4). In this paper, we assess if the ecological evidence supports the existence of a macrolide consumption threshold for the induction of macrolide resistance in *Streptococcus pneumoniae, Treponema pallidum* and *Mycoplasma genitalium*.

Ecological studies are justified and important in this field as the pathway between consumption and AMR operates at both individual and population levels. Lipsitch et al. outlined four mechanisms by which antibiotic treatment could select for resistance at a population-level (5). As an example, treatment with macrolides is likely to eradicate susceptible streptococci from an individual leading to the individual both being less likely to transmit susceptible streptococci to others and being at a higher risk of colonization by resistant streptococci (5, 6). Both these effects may be easier to detect at a population-than an individual-level (5, 6).

The relative importance of bystander selection to the emergence of AMR varies by bacterial species according to factors such as the proportion of the time that the bacteria circulate in a population as an asymptomatic colonizer (7). All three organisms chosen for the current analysis are asymptomatic for the majority of the time they circulate in a population (7-10). In addition, an ecological association has been found between population level consumption of macrolides and macrolide resistance for each of these organisms. For streptococci, for example, previous papers have found strong associations between macrolide consumption and AMR in *S. pneumoniae* (11-13) and *group A Streptococcus* (12). This association has been found to apply at national (12, 14) and sub-national levels (11, 15). Similar results have been found for *T. pallidum* and *M. genitalium* (8, 16). These analyses all used linear regression to assess the association between consumption and resistance and did not assess for the existence of a threshold effect. We were unable to find a study that evaluated if there was evidence of antimicrobial consumption thresholds in any of these species.

## Methods

### Data

#### Streptococcus pneumoniae

##### Antimicrobial Resistance Data

The *S. pneumoniae* susceptibility data from 2005 to 2018 was taken from the European Centre for Disease Prevention and Control (ECDC) Surveillance Atlas which reports resistance prevalence estimates from the European Antimicrobial Resistance Surveillance Network (EARS-Net) - the EU’s main surveillance system for AMR in bacteria that cause serious infections. All 28 EU Member States and two EEA countries (Iceland and Norway) participate in EARS-Net, but one country (Greece) does not report data on *S. pneumoniae*. Only data from invasive (blood and cerebrospinal fluid) isolates are included in EARS-Net. This is done to limit biases that may emerge if isolates from all anatomical sites were included. Further details can be found in the EARS-Net reporting protocol (17). The system depends on national network representatives in each participating country, reporting their locally tested susceptibility data to The European Surveillance System on an annual basis. The data for macrolide resistance reports the prevalence of resistance (as a percent) to erythromycin, clarithromycin or azithromycin. The details of the survey methodology and susceptibility testing are available at https://www.ecdc.europa.eu/en/surveillance-atlas-infectious-diseases. For each country, we calculated the median prevalence of macrolide resistance in the years 2005 to 2018.

##### Antimicrobial consumption data

Data from the European Surveillance of Antimicrobial Consumption (ESAC) were used as a measure of national general population-level antimicrobial drug consumption (18, 19). ESAC provides open access to the data collected on antimicrobial use in ambulatory care and hospital care in 30 European countries (18, 19). ESAC reports antimicrobial consumption as the number of defined daily doses (DDD) per 1000 inhabitants per day (DID) following the World Health Organization guidelines (19, 20). One DDD is defined as the average maintenance dose per day for a drug used in its main indication for adults (19, 20). We used ATC group J01F (macrolides, lincosamides and streptogramins), which is how ESAC reports macrolide consumption. Data was available from 1998 to 2018.

#### Treponema pallidum

The data for the prevalence of genotypic (A2058G or A2059G, *Escherichia coli* numbering) macrolide resistance in *T. pallidum* were extracted from a global country-level study that used linear regression to assess the association between macrolide consumption and resistance in *T. pallidum* (8). The national prevalence of genotypic macrolide resistance was based on a literature review (8). Where more than one prevalence estimate was available per country, the peak prevalence of resistance was used. The macrolide consumption data, expressed in DID, (for the year prior to that which provided the resistance prevalence) was taken from MIDAS Quantum database of the marketing research company IQVIA (IQVIA, Danbury, CT). IQVIA uses national sample surveys that are performed by pharmaceutical sales distribution channels to estimate antimicrobial consumption from the volume of antibiotics sold in retail and hospital pharmacies.

#### Mycoplasma genitalium

The data for both AMR and macrolide consumption were taken from a global study that compared macrolide consumption and resistance in *M. genitalium* (16). The prevalence of genotypic resistance to macrolides (mutations at positions 2058 or 2059 in 23S rRNA) used in this study was the country prevalence of macrolide resistance estimated by a systematic review of the prevalence of macrolide and fluoroquinolone resistance in *M. genitalium* (9). The macrolide consumption data used in this analysis was taken from the year prior to the resistance prevalence estimate (16). The macrolide consumption data, in DID, was obtained from the same IQVIA source described above.

### Timing of emergence of macrolide resistance in relation to total macrolide and azithromycin consumption

A number of studies have suggested that the widespread use of the long acting macrolide, azithromycin has played an important role in the genesis of macrolide resistance in *S. pneumoniae* and *T. pallidum* (10, 21, 22). To illustrate the putative association between total macrolide/azithromycin consumption and macrolide resistance we plotted the emergence of AMR by year in *T. pallidum (23, 24), M. genitalium* (9) and *S. pneumoniae* (10) in the United States in relation to the increase in sales of azithromycin in this population (sales were used as a proxy for consumption levels) (10) and total macrolide consumption. The data for azithromycin sales were taken from a paper that obtained annual sales from the manufacturer’s (Pfizer) annual financial reports (10). Sales data was available till 2005 when the azithromycin patent expired in the USA. Total macrolide consumption in DID was obtained from the IQVIA database described above. Data was available for the years 2000 to 2015. The resistance prevalence estimates for *T. pallidum* were taken from two STI clinics (San Francisco and Seattle (22, 23), for *M. genitalium* from the systematic review described above (9) and for *S. pneumoniae* from a paper that compiled resistance prevalence estimates from a literature review of the topic (10).

#### Analyses

We used an unvalidated but novel and simple method to define a putative macrolide consumption threshold for AMR. The US Centers for Disease Control and Prevention, the World Health Organization and others have recommended changing treatment protocols for *Neisseria gonorrhoeae* once the prevalence of AMR exceeds 5% (25, 26). Although somewhat arbitrary, we used >5% as our threshold of high prevalence of resistance. For each bacterial species, we then defined the *macrolide-5% threshold* as the highest median level of macrolide consumption of countries with a median resistance prevalence ≤5% for *S. pneumoniae* and *M. genitalium* and the peak resistance prevalence 5% for *T. pallidum*.

We compared differences in macrolide consumption between groups of countries using the Wilcoxon rank sum test. The statistical analyses were performed in Stata 16.0. A P-value of <0.05 was regarded as significant.

### Ethics approval

This analysis involved ecological analyses of public access data, and thus, no Ethics approval was necessary.

## Results

In the countries with available macrolide resistance estimates, the consumption of macrolides varied considerably. In the European Surveillance of Antimicrobial Consumption dataset, macrolide consumption was relatively stable between 1998 and 2018 - with the notable exceptions of Bulgaria, Ireland, France and Poland (Fig. 1). The median country macrolide consumptions varied between 0.8 DID in Sweden, and 4.9 DID in Italy (median 2.9 DID, IQR 2.2-3.4). Likewise, in the global IQVIA dataset, macrolide consumption varied considerably (Fig. 2).

**Figure 1.**
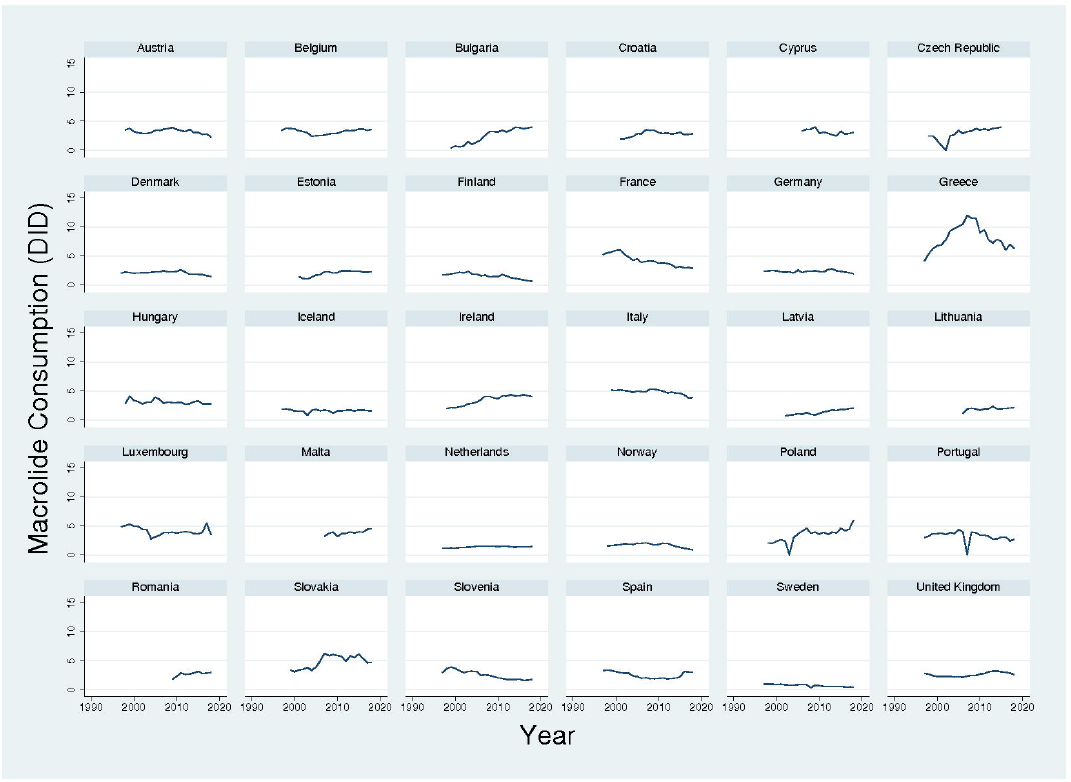
Macrolide consumption in 30 European countries, 1997-2018, according to European Surveillance of Antimicrobial Consumption. Data is in defined daily doses/1000 inhabitants/day (DID).

**Figure 2.**
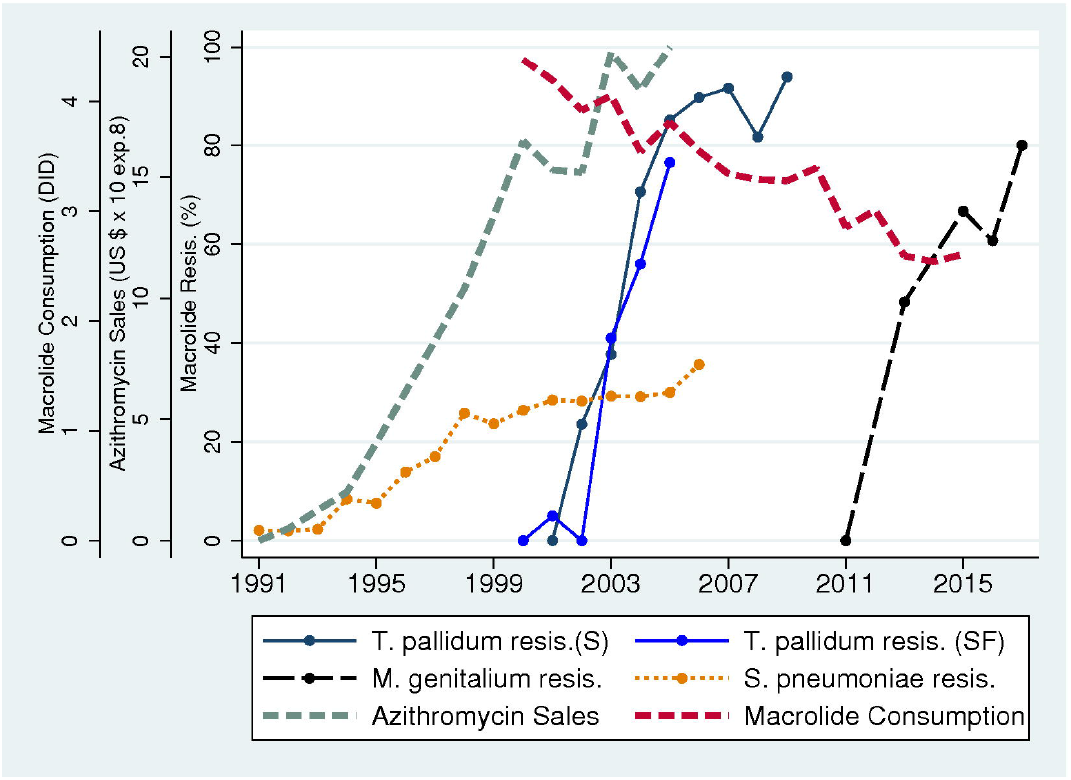
Country-level associations between macrolide consumption (in median number of defined daily doses/1000 individuals/day [DID]) and antimicrobial resistance in *Streptococcus pneumoniae* (a), *Treponema pallidum* (b) and *Mycoplasma genitalium* (c). In each graph, the blue horizontal line depicts 5% macrolide resistance.

Large variations were also evident in the prevalence of macrolide resistance to *M. genitalium, T. pallidum* and *S. pneumoniae* (Fig. 2).

### Macrolide-5% threshold in S. pneumoniae

The prevalence of macrolide resistance was ≤5% in 6 countries - Sweden, the Netherlands, Latvia, Norway, Denmark and Estonia (Fig. 2). The highest macrolide consumption in these countries was 2.3 DID in Estonia (median 1.6, IQR 1.3-2.2). The prevalence of macrolide resistance was lower in countries that consumed less versus more macrolides than this threshold – median 5.2% (IQR 4.5-12.7%) and median 20.1% (IQR 15.6-26.2%), respectively; P<0.001.

### Macrolide-5% threshold in T. pallidum

The prevalence of macrolide resistance in *T. pallidum* was ≤5% in 5 countries (Peru, Madagascar, South Africa, Taiwan and Russia). Of these countries, macrolide consumption was highest in Taiwan (1.8 DID). The prevalence of macrolide resistance was considerably lower in countries that consumed less versus those that consumed more macrolides than this threshold – median 1.2%, (IQR 0-14.3%) and median 84%, (IQR 66.7-100%), respectively; P=0.011.

### Macrolide-5% threshold in M. genitalium

Macrolide resistance was ≤5% in 2 countries (Kenya and Russia). Macrolide consumption data was not available for Kenya and was 1.3 DID in Russia. The prevalence of macrolide resistance was higher in countries above this threshold (median 41%, IQR 12.6-61.3%) than Russia (3.7%) but this difference was not statistically significant (P=0.137).

## Timing of emergence of macrolide resistance in relation to azithromycin consumption

Azithromycin sales in the United States increased rapidly from close to zero dollars in 1991 to over $2 billion in 2005 (Fig. 3). This increase was followed approximately three years later by an increase in macrolide resistance in *S. pneumoniae* from close to zero to above 30%. The rapid increases in macrolide resistance in *T. pallidum* and *M. genitalium*, however, followed approximately 10 and 20 years after the increase in azithromycin consumption. Total macrolide consumption was high in 2000 (4.4 DID) and, whilst it decreased over time (2.6 DID in 2015), it remained high compared to consumption in countries such as Sweden (0.6 DID in 2015).

**Figure 3.**
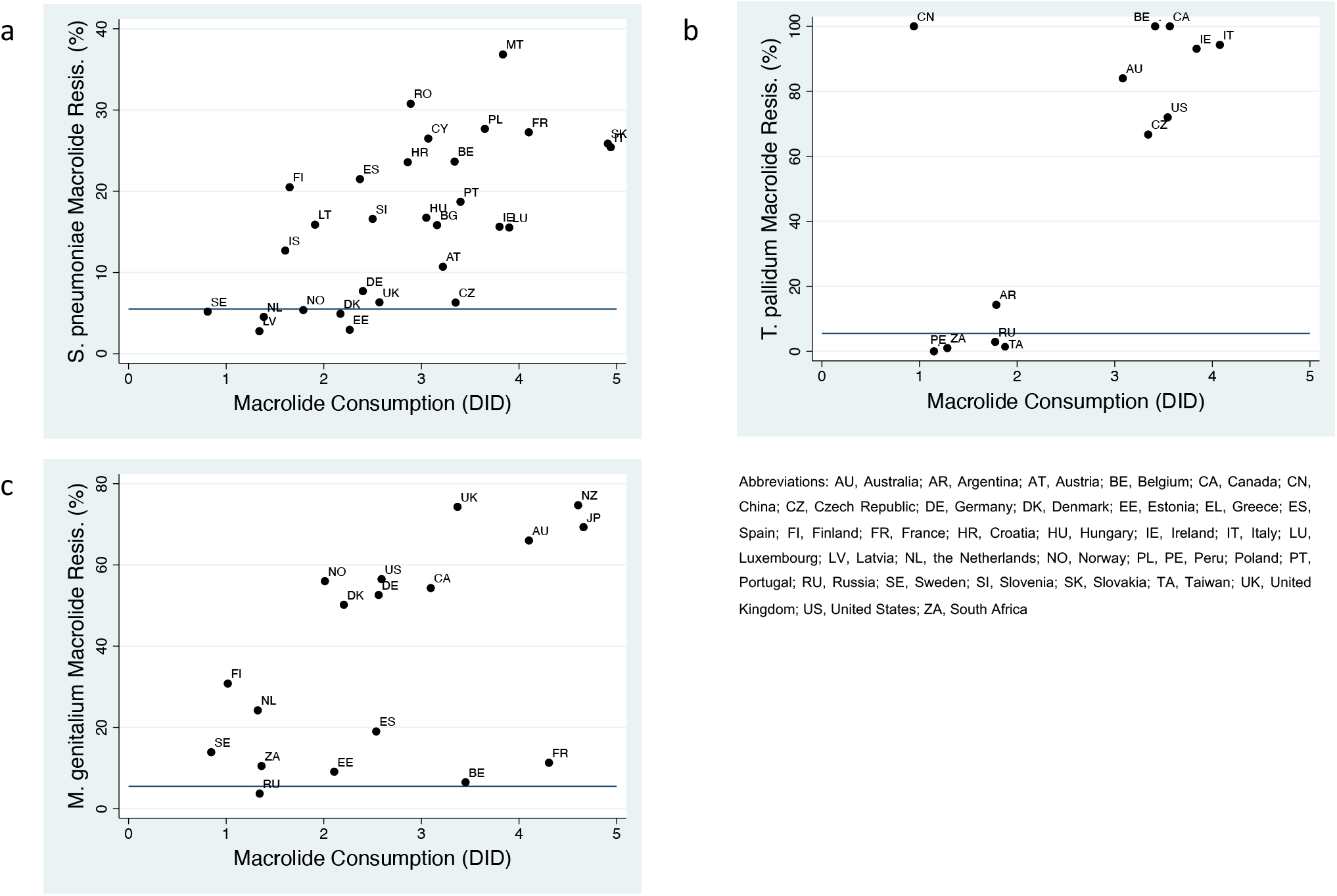
Temporal relationship between macrolide consumption and resistance in the United States. The emergence of macrolide resistance in *Streptococcus pneumoniae* in the United States followed a few years after the steep increase in azithromycin sales (US $ ⨯10^8^). The emergence and spread of macrolide resistance in *Treponema pallidum* and *Mycoplasma genitalium* in the United States followed one to two decades later. Total macrolide consumption (in defined daily doses/1000 inhabitants/day – DID) declined slightly after 2000 but remained high compared to other countries.

## Discussion

The application of our *macrolide-5%* methodology produced evidence of a macrolide consumption threshold of 1.3 DID for *M. genitalium*, 1.8 DID for *T. pallidum* and 2.3 DID for *S. pneumoniae*.

There are a number of caveats to this conclusion, not the least of which is that we have only investigated the impact of a single variable on the genesis of AMR. The emergence and spread of AMR is dependent on multiple factors acting at different levels (1, 2, 27, 28). These factors include homologous and heterologous class antimicrobial consumption (individual and population level), vaccination, travel and sanitation (1, 2, 8, 27-29). The relative contributions of specific risk factors (such as population level macrolide consumption) is also likely to vary between bacterial species according to factors such as the mechanisms whereby they acquire macrolide resistance as well as their susceptibility to bystander selection (7, 29, 30). This is evident if we consider the differences in the timing of the emergence of macrolide resistance in the three organisms in the USA. If the effect of population level macrolide consumption on AMR was uniform, then we would expect macrolide resistance to emerge concurrently. A number of factors may play a role in explaining differences in the timing of resistance emerging. The prevalence of *S. pneumoniae* is typically higher than that of *T. pallidum* and *M. genitalium* which may have placed it under a greater selection pressure from population level consumption of macrolides (7). The incidence of *T. pallidum* in the USA was low in the 1980s and early 1990s and only began to increase in the late 1990s which may have meant that *T. pallidum* would only have been placed under a selection pressure from macrolides consumed for other purposes in the late 1990s (31). The data from the Seattle cohort supports this conclusion. Macrolides were not used to treat syphilis in this center, but a high proportion of clients (27%) had received macrolides for other indications in the 12 months prior to their syphilis diagnosis and self-reported macrolide use was associated with macrolide resistance in *T. pallidum* (22). The delayed increase in macrolide resistance in *M. genitalium* may be explained by changes in treatment guidelines for STIs such as the use of azithromycin in combination with ceftriaxone as first line treatment for N. gonorrhoeae in 2010 (26). A complementary explanation is that increases in screening and treatment of STIs that can be treated with azithromycin such as chlamydia and *M. genitalium* played a role (32, 33).

A number of other factors may explain the differences in the association between macrolide consumption and resistance in the three species. Differences in the fitness costs of resistance associated mutations may be important. Variations in the biological pathways to macrolide resistance between the three species also likely play an important role. *S. pneumoniae* can take up resistance conferring sections of DNA (such as *erm*-type methylases and *mef*-type efflux pumps) via transformation and conjugation (29, 34). This process has been shown to have played an important role in its acquisition of resistance to macrolides (34). *M. genitalium* and *T. pallidum* do not have this ability and the acquisition of resistance typically results from chromosomal mutations in the gene coding for the 23s RNA (8, 9, 22).

There are also a number of countries with low reported consumption of macrolides but a high prevalence of macrolide resistance. The high prevalence of macrolide resistance in *T. pallidum* in China is an example (Fig. 2). These examples could be explained by high macrolide consumption in local populations with high prevalence of circulating *T. pallidum* (such as core-groups) or the acquisition of resistant strains via travel from high consumption countries (8). The very high prevalence of macrolide resistance in other bacterial species, as well as *T. pallidum* in China could also be explained by a false low estimate of macrolide consumption in this population (8, 30). It is possible that countries with ≤5% macrolide resistance in these bacterial species will still cross this resistance threshold. Evidence along these lines is available for the two countries (France and Belgium) with low macrolide resistance in *M. genitalium* (6.5% and 11.3%, respectively) but high macrolide consumption (Fig. 2). The resistance prevalence estimates from both countries were derived from relatively old studies used in the systematic review (9). More recent studies have found a much higher prevalence of macrolide resistance in both countries (74% and 58%, respectively) (35, 36). These limitations mean that the results should be viewed with caution and require further validation. The way that travel can disseminate AMR may also mean that for certain bacterial species, a threshold may only be effective if globally applied (37).

Despite these limitations, the study may provide guidance to health planners. For example, there is considerable debate in the literature as to the net benefits and risks of screening men who have sex with men in pre-exposure prophylaxis (PrEP) cohorts for gonorrhoea and chlamydia (32, 33, 38, 39). PrEP guidelines recommend up to 3-monthly screening (38), but this has been shown to result in the consumption of approximately 12.1 DID of macrolides (33). In the absence of a rough consumption-resistance threshold, it is difficult for planners to know how to interpret this 12.1 DID figure. If we use the macrolide-5% thresholds outlined here, then a consumption of 12.1 DID macrolides could be seen as being between 5- and 9-fold higher than the resistance thresholds for *S. pneumoniae, M. genitalium* and *T. pallidum* that were found in this study. In the absence of any evidence from randomized controlled trials showing any benefit from screening (38), planners may use this information to reevaluate the need/frequency of chlamydia/gonorrhoea screening in this population. They could also use this information to justify cutting back screening to the PrEP clients. One option would be to visualize the macrolide consumption as a ‘macrolide consumption footprint’ which could be compared to the threshold footprint (4).

Whilst our analysis has not evaluated the extent to which macrolide resistance is reversible, longitudinal studies in Finland have found that a decline in macrolide consumption was followed by a significant decline in the frequency of erythromycin resistance in group A streptococcal isolates (17 to 9%) (40). Likewise, studies in Scotland have found that reductions in macrolide consumption were temporally associated with declines in the incidence of *Clostridium difficile* (41) and methicillin resistant *Staphylococcus aureus* infections (42). Taken in conjunction with this evidence, our study provides further motivation for macrolide/antimicrobial stewardship campaigns that strive to reduce macrolide consumption to levels at least below 2 DID, but if the burden of infection allows, preferably to under 1 DID as achieved in countries such as Sweden.

## Data Availability

All data is available as described in the methods section

## Acknowledgements

We would like to thank the ECDC for the data provided on their Surveillance Atlas of Infectious Diseases

## Authors’ contributions

CK conceptualized the study. CK was responsible for the acquisition, analysis and interpretation of data. CK, SMB and CVD contributed to the first draft and read and approved the final draft.

## Consent for publication

Not applicable

## Data availability

The data we used is publicly available from the papers we refer to and the ECDC data is available from: https://atlas.ecdc.europa.eu/

## Funding

No specific funding was received for this work.

## Competing interests

None to declare. All the authors declare that they have no conflicts of interest.

## Notes

### Competing Interest Statement

The authors have declared no competing interest.

### Funding Statement

No funding was received for this study

### Author Declarations

Institute of Tropical Medicine IRB

